# Patient Experiences of Treatment Deferral for Evaluation and Monitoring after a Trace Xpert Ultra Result

**DOI:** 10.1101/2025.01.22.25320963

**Authors:** Caitlin Visek, James Mukiibi, Mariam Nantale, Annet Nalutaaya, Patrick Biché, Joowhan Sung, Francis Kayondo, Joab Akampurira, Michael Mukiibi, Rogers Kiyonga, Achilles Katamba, Emily A. Kendall

## Abstract

**Background:** A “trace” result from the Xpert Ultra molecular TB test indicates *Mycobacterium tuberculosis* DNA detection but may not always signify TB disease. Little is known about the experiences of individuals with trace results who are not immediately treated.

**Methods:** We enrolled Ugandan adults and adolescents with trace Xpert Ultra sputum results, plus positive and negative controls, from community-screening and clinic settings. After a comprehensive TB evaluation, participants not recommended to start treatment immediately were closely monitored with interval reassessments. Surveys captured participants’ perceptions and preferences related to their uncertain TB status at baseline and during follow-up.

**Results:** A total of 321 people with trace sputum (PWTS), 236 positive controls, and 279 negative controls were enrolled. A minority of PWTS thought they were highly likely to have or develop TB, and most reported low associated anxiety initially (258/321, 80%) and during follow-up. While about half (172/321, 54%) would have favored treatment at baseline if not in the study, those who remained untreated were less inclined toward treatment at six months. Participants generally preferred sensitive diagnostic tests, even with frequent false positives.

**Conclusions:** Deferring treatment for PWTS, with sufficient testing and monitoring, is acceptable to most patients.

## INTRODUCTION

Xpert MTB/RIF Ultra (“Xpert Ultra”; Cepheid) is a rapid molecular test widely used for diagnosing tuberculosis (TB). Compared to the prior generation assay (Xpert MTB/RIF), Xpert Ultra is able to detect *Mycobacterium tuberculosis* (MTB) in sputum at lower bacillary burdens.^1^ However, results in the lowest category of positivity, termed “trace,” are often associated with negative sputum cultures and have been interpreted as false positive results.^2-5^ Lacking reliable procedures for distinguishing between true- and false-positive trace results, current practice and World Health Organization guidance favor providing TB treatment to most people with trace sputum (PWTS).^6^ Still, for some PWTS who have mild or resolving illness (or who are asymptomatic) without other known evidence of TB disease, forgoing immediate treatment in favor of monitoring and repeat testing may be an acceptable alternative.

Little is known about how patients might experience decisions to forgo immediate treatment or discussions about diagnostic uncertainty. In other clinical contexts, people with TB can experience emotional distress related to fear of stigma, disease transmission, and death.^7-9^ If patients are recommended to undergo monitoring rather than upfront treatment, they must contend with uncertainty regarding their TB status as well as the time and inconvenience of additional diagnostic evaluation. To better understand this experience, we surveyed cohorts of people who received trace results from an Xpert Ultra test during either TB diagnostic evaluation or community-based TB screening. We asked about their perception of their TB status, any anxiety related to the associated uncertainty, and their experiences and preferences regarding subsequent TB testing and management. We compared their testing preferences to those of people with more definitively positive or negative Xpert Ultra results, and we repeated the survey over time to assess changes in attitudes toward trace results and treatment deferral.

## METHODS

### Study design and population

People with trace-positive sputum, and individuals with positive and negative Xpert results who served as controls, were identified through both community-based screening and clinic-based diagnostic evaluations in Kampala, Uganda between February 2021 and October 2024. Community-based screening occurred in high-risk areas without preceding screening tests, while clinic-based testing was typically symptom-driven. In both settings, expectorated sputum samples were tested for tuberculosis with Xpert Ultra, and all individuals with trace results were contacted for enrollment. For most of the study period we also recruited controls: “negative controls” with negative Xpert results (matched approximately 1:1 with enrolled PWTS in each setting), and “positive controls” with positive results at a level greater than trace (enrolled universally from community screening where approximately half of all positive results were trace, and matched 1:1 in the clinic setting). Eligible participants were at least 15 years old and were not on TB treatment at the time of testing or (for clinic-recruited participants) within the preceding 90 days. For PWTS, the informed consent process explained that the trace result “does not provide a clear answer about whether or not you have TB,” that it might indicate TB with a low bacterial burden or might be falsely positive, and that the study would include additional testing and follow-up to determine TB status and treatment needs.

All consenting participants underwent a comprehensive clinical, laboratory, and radiographic evaluation at baseline (including standardized interview, chest x-ray (CXR) and CT, medical history and physical, and laboratory testing including repeat sputum Xpert Ultra, sputum culture, TB immunoreactivity [QuantiFERON] testing, TB tongue swab testing,^10^ HIV testing, and serum C-reactive protein (CRP) measurement, as well as urine lipoarabinomannan (LAM) and CD4 count measurement for HIV-positive participants.

At baseline and on an ongoing basis, an independent physician review panel reviewed available clinical and laboratory data from PWTS and recommended treatment initiation whenever they judged that a given patient was highly likely to have TB or at high risk for harm from continued treatment deferral. Study-unaffiliated clinicians who cared for study participants could also initiate treatment at any time. For the current analysis, we consider a TB diagnosis to be microbiologically confirmed if the participant had either a sputum culture positive for MTB or a repeat Xpert Ultra result that was positive at a level greater than trace. Participants not recommended for treatment after their baseline evaluations underwent longitudinal monitoring with periodic repeat clinical, laboratory, and imaging evaluations (initial follow-up visits scheduled at one, three, and six months). Negative-control participants were reevaluated less frequently (first follow-up visit at six months). Positive-control participants were referred for TB treatment and had no further study follow-up.

The study was approved by the Makerere University School of Public Health Research and Ethics Committee and the Johns Hopkins Medicine Institutional Review Board.

### Patient experience survey

Beginning on September 30, 2021, a structured questionnaire about subjective experiences and preferences was administered to participants at study enrollment and at one- and six-month follow-up visits. Questions asking participants to rate aspects of their experience used three-to six-level Likert scales.

The baseline questionnaire was administered after participants had been informed of their initial screening or diagnostic Xpert results and enrolled into the study. PWTS were asked how anxious they felt about the possibility of having or developing TB and whether they would be inclined to seek out TB treatment if they were not being offered additional testing and monitoring via the study. Those enrolled from the community were also asked to rate whether, considering everything they knew at the time, they believed themselves to currently have TB, and whether they thought themselves likely to develop TB in the future.

The baseline survey also included a series of vignette-based questions aimed at evaluating tradeoffs between sensitivity and specificity. All participants were asked to choose between alternative TB tests to recommend to their friends and family. One hypothetical test (representing high sensitivity but poor specificity) was labeled as “too strong” and described as correctly identifying everyone with TB but producing one false-positive result for every TB case correctly identified. The other (representing low sensitivity but excellent specificity) was labeled as “too weak” and described as generating no false-positive results but misclassifying one out of every two people who truly have TB as TB-negative. Participants who chose the initial version of the “too strong” test were also presented with a follow-up scenario in which the number of false positives per true positive increased five-fold.

At the one-month (for PWTS) and six-month (for PWTS and negative controls) follow-up visits, participants who remained off TB treatment were surveyed again. They were asked to rate how inconvenient or unpleasant they had found completing each of the diagnostic tests they completed previously, and how valuable they considered each of the tests. Survey questions about anxiety, treatment inclination, and self-perception of TB status were also repeated.

### Statistical analysis

All analyses were performed with R Statistical Software version 4.3.1 (R Core Team, 2023) and RStudio version 2023.06.1+524 (Posit). Categorial variables were compared using Pearson’s chi-squared test or Fisher’s exact test with a significance level of 0.05.

## RESULTS

A total of 836 participants were enrolled during the period included in this analysis: 378 recruited from community-based screening events and 458 from clinic settings (**Figure 1**). These included 321 participants with a recent trace Xpert Ultra result (enrolled as PWTS), 236 positive controls, and 279 negative controls. Of the 321 enrolled PWTS, 101 (31%) started treatment prior to the one-month follow up visit, and 140 (44%) by the six-month visit.

**Figure 1.**
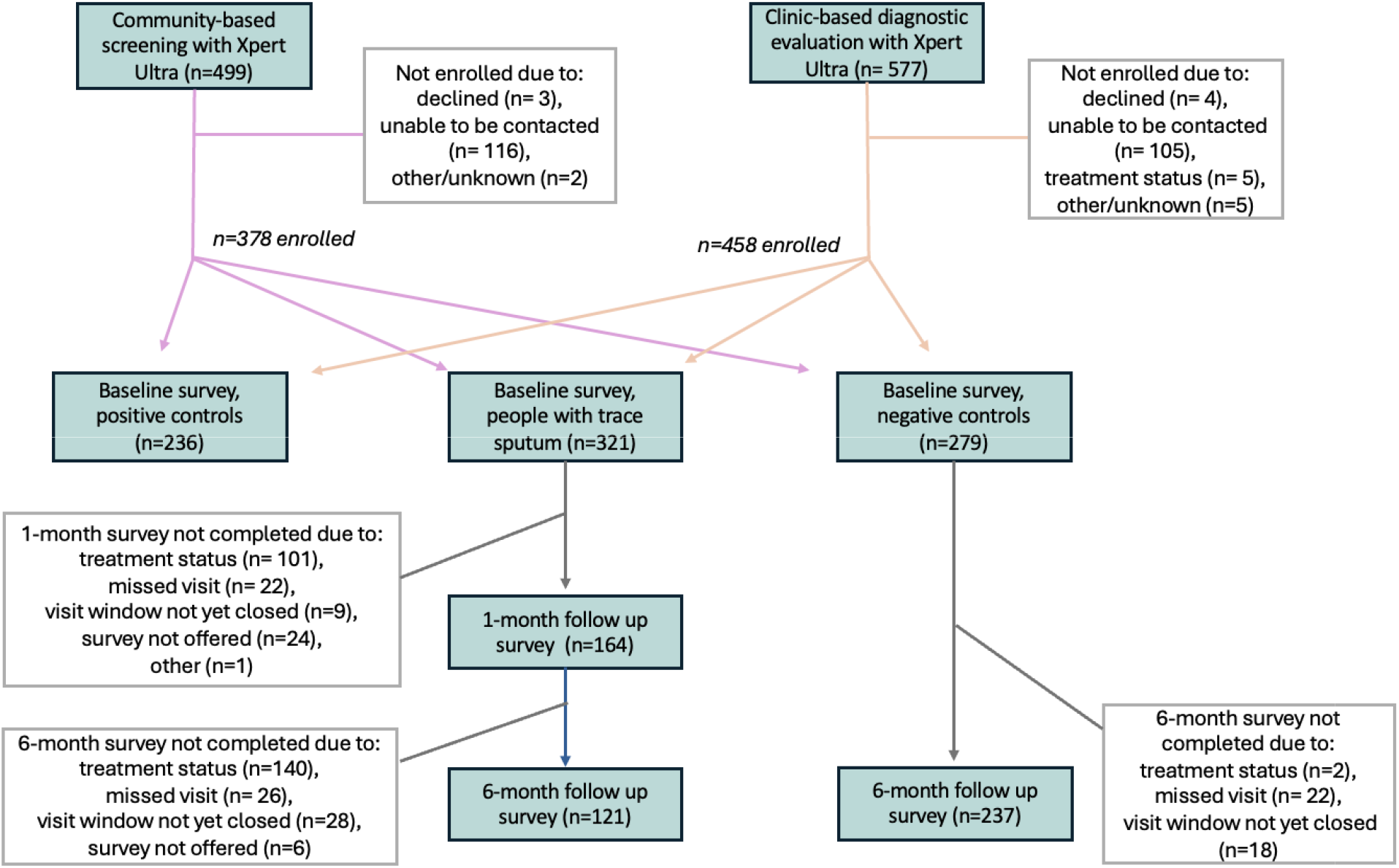
Overview of study population

At baseline, a majority of PWTS (258/321, 80%) reported little to no anxiety at baseline about the possibility that they could have or develop TB, while 47/321 (15%) reported moderate anxiety and 16/321 (5%) reported feeling very anxious. Anxiety was lowest in the community setting (**Table 1**). About half of participants in both settings reported that they would be inclined to start TB treatment were they not enrolled in the study. For PTWS enrolled from the community setting, who were asked about perceived likelihoods of having or developing TB, a minority perceived themselves as very likely to have TB currently (28/129, 22%) or develop it in the future (30/129, 23%). Higher perceived likelihoods of having TB currently or of developing TB in the future (among community PTWS) and greater inclination toward treatment (among all PWTS) were significantly associated with being started on treatment by six months (p-values 0.02, 0.02, and 0.02, respectively).

**Table 1.** Anxiety, treatment preferences, and interpretations of TB status reported at baseline by people with a trace sputum result, by source of enrollment and by treatment status.

|  | Enrollment Setting |  | Treatment Status |  |
| --- | --- | --- | --- | --- |
|  | Community<br>N= 129 | Clinic<br>N= 192 | Not treated by 6m<br>N= 181 | Treated by 6m<br>N= 140 |
| Reported moderate to high anxiety about possibility of having or developing TB, N (% of column) | 14 (11%) | 49 (26%) | 34 (19%) | 29 (21%) |
| Reported inclination towards starting treatment had they not been in a research study, N (% of column) | 67 (52%) | 105 (55%) | 87 (48%) | 85 (61%) |
| Perceived themselves to probably or certainly/almost certainly have TB now, N (% of column)* | 28 (22%) | Not asked | 11/77 (14%) | 17/52 (33%) |
| Perceived themselves as probable or certain/almost certain to develop TB at some point in the future, N (% of column)* | 30 (23%) | Not asked | 11/55 (20%) | 19/43 (44%) |

\*Not asked of participants enrolled from clinic settings

One month after enrollment, 101/321 (31%) PWTS had started TB treatment, including 10/16 (63%) of those who were very anxious at baseline (**Figure 2**). Of those 10, only 3 had microbiological confirmation of disease (compared to 59% of those started on treatment overall). Most who remained off treatment reported either an unchanged (96/164, 59%) or a decreased (40/164, 24%) level of anxiety at one-month follow-up, while 28/164 (17%) reported an increase. By six months, two-thirds of participants who remained off treatment reported no anxiety (82/121, 68%), no participants reported feeling very anxious, and only 14 (14%) of the 104 participants who completed both follow up surveys reported a higher level of anxiety at six months than at one month (**Figure 2**).

**Figure 2.**
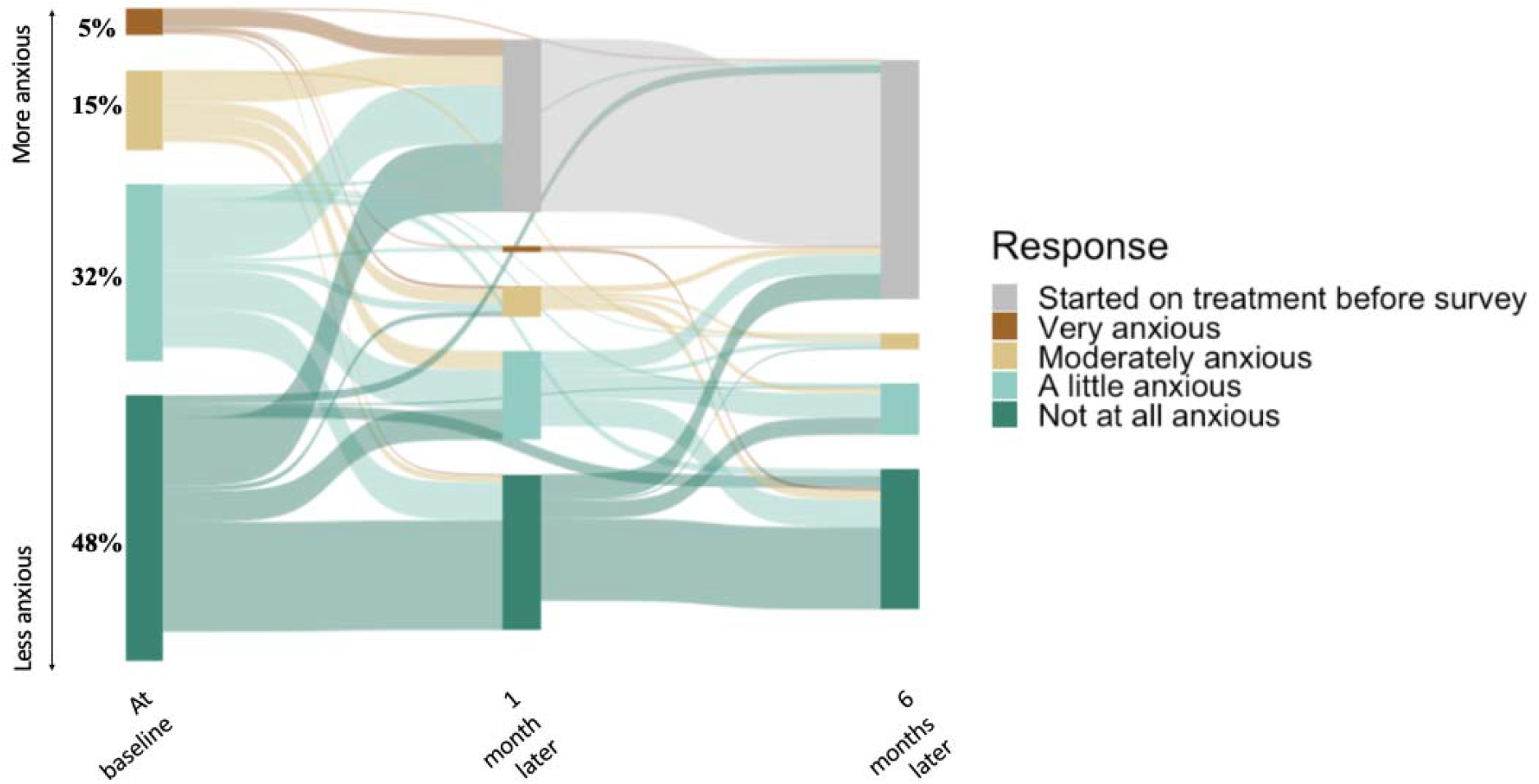
Reported anxiety associated with a trace Ultra result, immediately after receiving the trace result and over 6 months of follow up

When asked whether they would be inclined to pursue TB treatment if they were not being offered monitoring and repeat testing as part of the study, a slight majority of PWTS expressed an inclination toward treatment at baseline (172/321, 54%). Most of those who remained untreated continued to report the same treatment inclination at one month (120/164, 73%) and six months (82/121, 68%) as at baseline, but participants whose opinion changed from baseline to six months were more likely to change toward not wanting TB treatment (31/39, 71%) than toward favoring TB treatment (8/39, 21%; p-value < 0.01) (**Figure 3)**. By six months, only 40/121 (33%) favored treatment.

**Figure 3.**
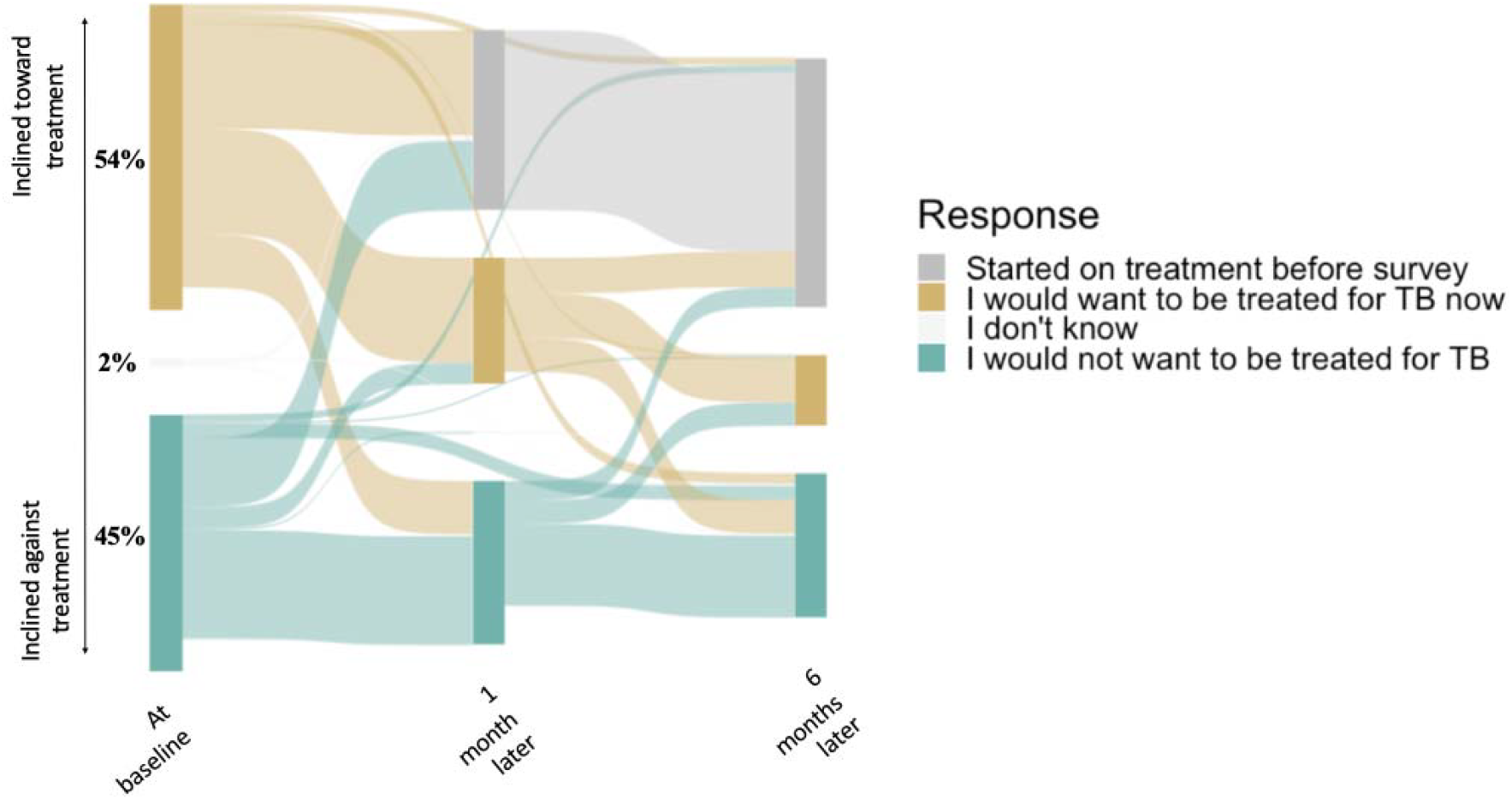
Self-reported inclination toward treatment, had participants not been part of a research study, immediately after receiving a trace Ultra result and over six months of subsequent follow up

When presented with choices between a more sensitive test and a more specific test, 59% of all participants (495/834) preferred the more sensitive option in the first scenario where this test’s positive predictive value (PPV) was 50% (**Table 2**). The inclination toward a more sensitive test was stronger among participants recruited from the community setting (70%) than from health facilities (51%; p-value < 0.01). Responses did not vary significantly by initial Xpert Ultra result. In the second scenario in which the “too strong” test had a lower (17%) PPV, the proportion preferring this more sensitive option decreased to 45%.

**Table 2.**
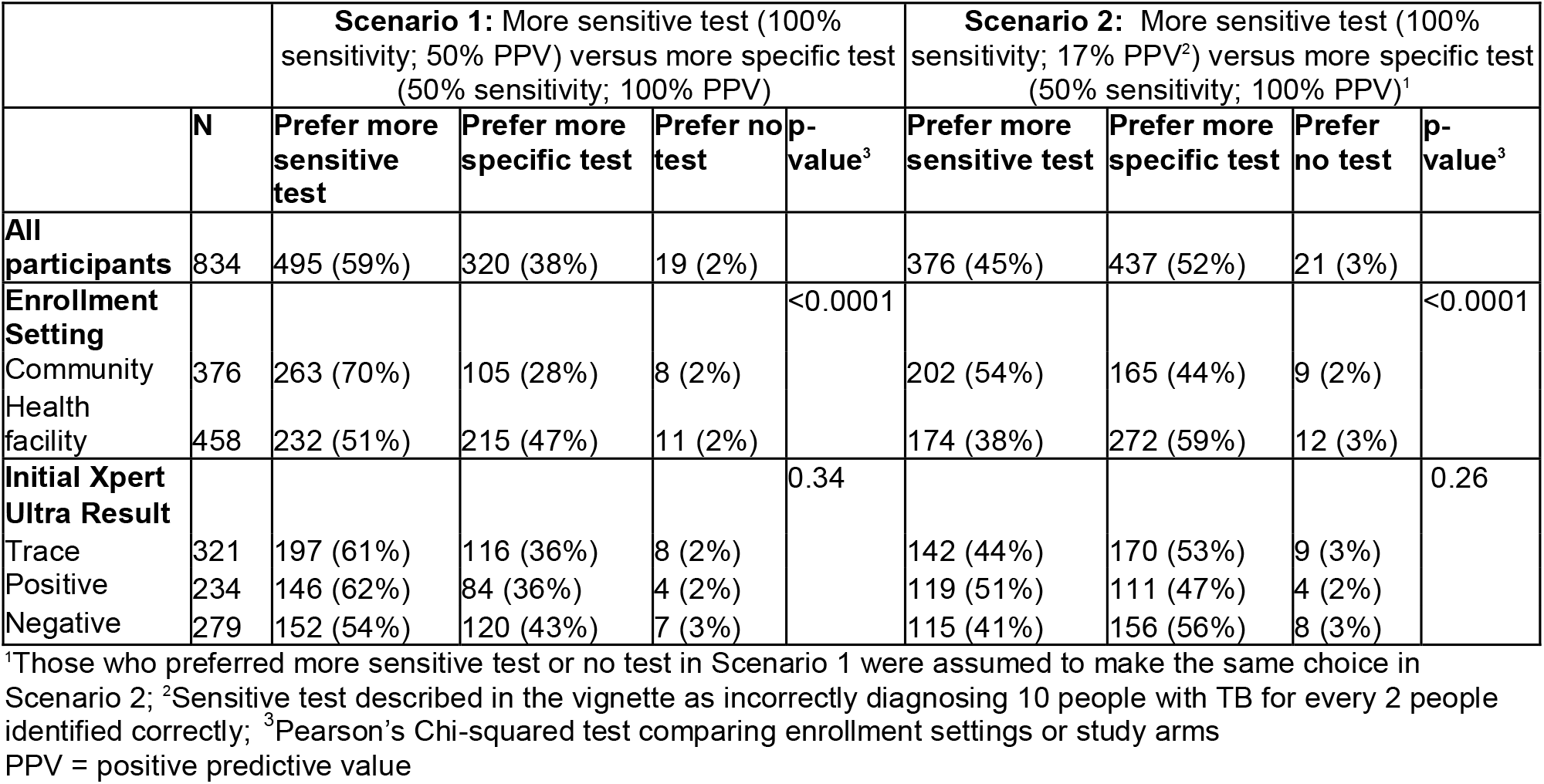
Participants’ preferences regarding TB test sensitivity versus specificity.

All diagnostic tests used to further evaluate PWTS were rated favorably by participants who had completed them, with nearly all participants reporting that they considered the tests valuable and did not mind completing them (**Supplemental Figure 1**).

## DISCUSSION

When managing trace sputum results with an approach of treatment deferral, further diagnostic evaluation, and close monitoring in the context of a research study, we found that half of surveyed PWTS perceived themselves as unlikely to have TB presently, a minority believed they were likely to develop TB in the future, and 80% reported no or little associated anxiety at the time of enrollment. Over the following six months, nearly 50% initiated treatment, but those who remained off treatment reported further reductions in anxiety and became less likely to favor initiating TB treatment. Participants also rated their experiences of completing multiple TB diagnostic procedures quite favorably. Altogether, our findings suggest that further diagnostic evaluation, with close monitoring off treatment for those who do not have other evidence of TB at baseline, is acceptable to the majority of PWTS.

For those PWTS least likely to have active TB, a monitoring approach may also minimize harms from overtreatment. PWTS identified through screening, for example, may be less likely to have bacteriologically confirmed TB; the few studies that have used Xpert Ultra as an initial screening test found positive sputum cultures in only 10-14% of people with trace results,^11, 12^ In surveys that screened with symptoms and/or CXR prior to Xpert Ultra testing, the prevalence of culture positivity among those with a trace result was higher though still under 55%.^5, 13, 14^ Patients with a history of prior TB are another population in whom trace results are relatively likely to be false-positive.^3, 4, 15^ An early report of longitudinal follow-up of PWTS not treated for TB suggests that some with negative baseline sputum cultures may still be at elevated risk to develop TB;^16^ however, if the lowest risk PWTS could be reliably identified, they may benefit from treatment deferral.

Ideally, decisions to treat a patient with a trace result would be based on an individual risk assessment followed by shared decision-making that takes both likely outcomes and patient preference into consideration. However, not only is the risk of TB in PWTS poorly understood, the ideal balance between potential harms of over- and undertreatment of possible TB is not well established. Surveys of clinicians suggest they may overweight harms of unnecessary TB treatment, requiring a higher probability threshold (between 20 and 60%) when making treatment decisions intuitively than when guided in systematic estimation of utilities (often leading to treating patients with <10% probability of TB).^17^ From the patient perspective, responses to our survey about tradeoffs between test sensitivity versus specificity suggest participants’ intuitive tolerance for overdiagnosis and overtreatment is at least as high as clinicians’, with a majority favoring a TB test with a PPV of 50% over one that would miss cases. Thus, deferring treatment for PWTS may only be appropriate when the probability of TB is particularly low, or when close follow-up can ensure that patients who have TB are highly likely to be identified and treated before they experience harm.

Even when further diagnostic testing and monitoring after a trace result would be acceptable to most patients and clinicians, the resources required may be unavailable. Diagnostic evaluations performed in this study, such as CT scans and repeated cultures, are often performed to assess uncertain TB status in high-resource settings but may be unavailable or prohibitively expensive in many places with a high burden of TB. Additionally, without dedicated personnel to follow up with participants, longitudinal monitoring may not be feasible. Importantly, we found that half of the PWTS who consented to the study would have preferred initiating TB treatment at baseline if they had not been enrolled in the study.

Our study has some limitations. Anxiety or desire to start TB treatment may have influenced patients’ care-seeking or symptom reporting, biasing which study participants remained untreated and eligible for follow-up surveys. Social desirability may have led patients to minimize any negative feelings, especially as the survey was administered by study staff. Participating in research that provided resources for further testing and follow-up may have alleviated anxiety about treatment deferral among PWTS. Finally, while our structured questionnaire facilitated summarization and comparison of responses across a large sample, more in-depth qualitative data would be needed to understand the range of patient experiences.

In summary, for patients with a trace Xpert Ultra result of uncertain clinical significance, deferring treatment decisions may be acceptable from the patient perspective when further diagnostic evaluation and monitoring can be offered. TB treatment decision-making for PWTS should consider individual TB risk, patient preferences, and available resources to support appropriate additional testing and monitoring. Further studies will be needed to optimize management of PWTS and assess the clinical impact of treatment decision deferral.

## Data Availability

All data produced in the present study are available upon reasonable request to the authors

## Acknowledgements

This work was supported by the US National Institutes of Health (R01HL153611 to EAK; T32AI007291 and K23AI185268 to JS) and the Bill and Melinda Gates Foundation (INV-042921 to EAK). The content is solely the responsibility of the authors and does not necessarily represent the official views of the funders.

## SUPPLEMENTARY DATA

**Figure S1.**
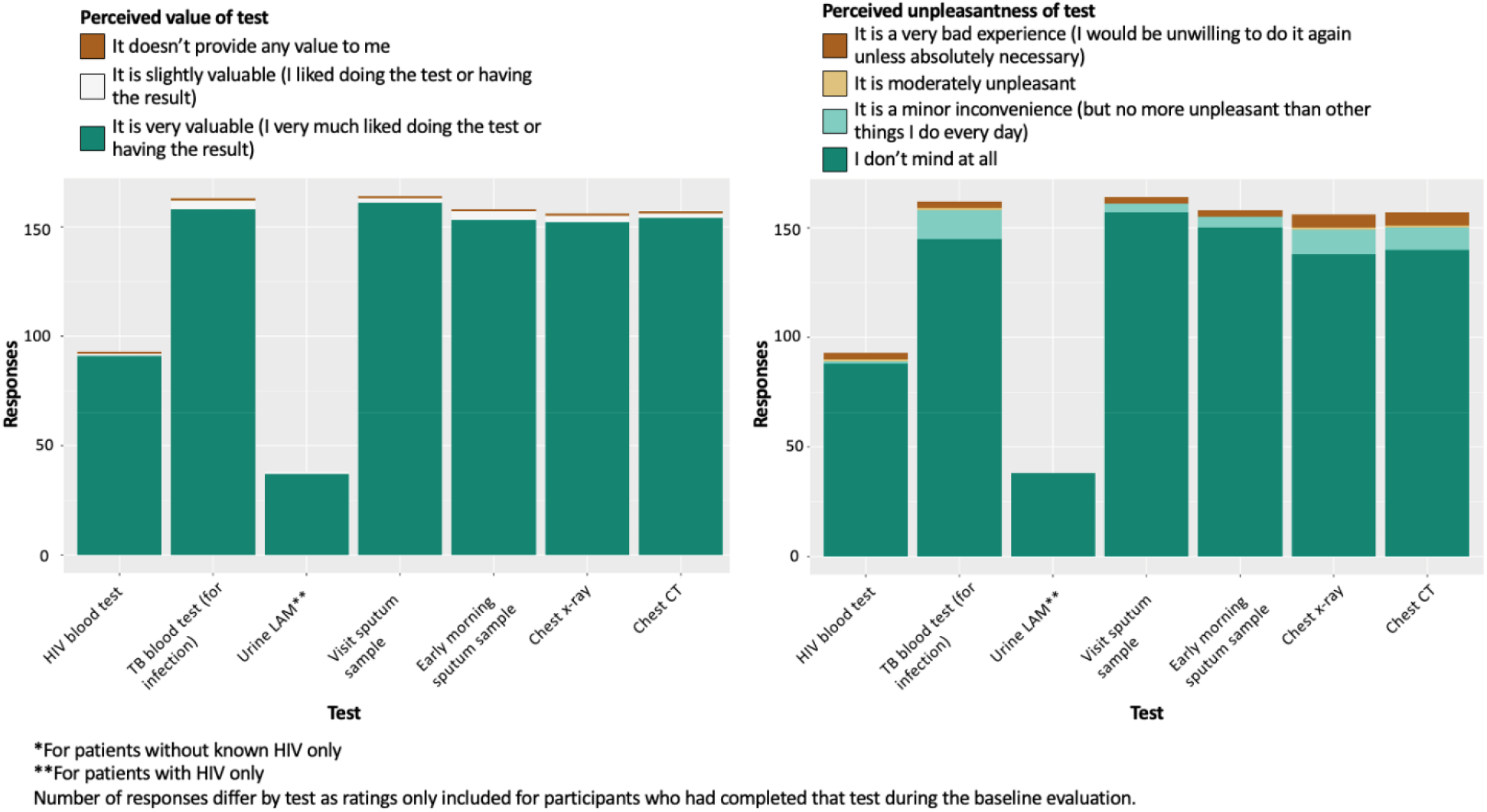
Ratings, by people with trace Ultra sputum results surveyed at the one-month follow-up visit, of the value and unpleasantness of each diagnostic test completed at baseline

